# Evaluating the impacts of tiered restrictions introduced in England in December 2020 on covid-19 hospitalisations: a synthetic control study

**DOI:** 10.1101/2024.02.28.24303487

**Authors:** Xingna Zhang, Daniel Hungerford, Mark A. Green, Marta García-Fiñana, Iain E. Buchan, Ben Barr

## Abstract

**Objectives:** To assess the impact of Tier 3 covid-19 restrictions implemented in December 2020 in England on covid-19 hospital admissions compared to Tier 2 restrictions, and its potential variations by neighbourhood deprivation levels and the prevalence of the Alpha variant (B.1.1.7).

**Design:** Observational study utilising a synthetic control approach. Comparison of changes in weekly hospitalisation rates in Tier 3 areas to a synthetic control group derived from Tier 2 areas.

**Setting:** England between 4^th^ October 2020 and 21^st^ February 2021.

**Participants:** 23 million people under Tier 3 restrictions, compared to a synthetic control group derived from 29 million people under Tier 2 restrictions.

**Interventions:** Implementation of Tier 3 covid-19 restrictions in designated areas on 7^th^ December 2020, with additional constraints on indoor and outdoor meetings and the hospitality sector compared to less stringent Tier 2 restrictions.

**Main Outcome Measures:** Weekly covid-19 related hospital admissions for neighbourhoods in England over a 12-week period following the interventions.

**Results:** The introduction of Tier 3 restrictions was associated with a 17% average reduction in hospital admissions compared to Tier 2 areas (95% CI 13% to 21%; 8158 (6286 to 9981) in total)). The effects were similar across different levels of neighbourhood deprivation and prevalence of the Alpha variant (B.1.1.7).

**Conclusions:** Regionally targeted Tier 3 restrictions in England had a moderate but significant effect on reducing hospitalisations. The impact did not exacerbate socioeconomic inequalities during the pandemic. Our findings suggest that regionally targeted restrictions can be effective in managing infectious diseases.

**SUMMARY BOXES:** *What is already known on this topic:*

— Previous studies of localised non-pharmaceutical interventions (NPIs) found that they could be effective in reducing covid-19 transmissions.
— covid-19 hospitalisation was a key indicator of healthcare resource dynamics, encompassing supply, demand, burden, and allocation, during the pandemic.
— There is a need for a detailed examination of the impact of specific localised restrictions in the UK, such as Tier 3 measures, on hospital admissions to inform targeted public health strategies.

*What this study adds:*

— This study found that additional localised restrictions on outdoor gatherings and in the hospitality sector were effective in mitigating hospital admissions during the pandemic.

*How this study might affect research, practice or policy:*

— This study provides evidence for future public health policies and preparedness strategies supporting the use of differential regional restrictions during pandemics.

## INTRODUCTION

Managing covid-19 hospitalisations was a major challenge during the pandemic.^1,2^ At the beginning of the pandemic, “flattening the curve” in hospitalisations was crucial in England.^3^ Nevertheless, England was one of the most severely affected countries, with some of the globally highest rates of covid-19 hospitalisations and deaths between late 2020 and early 2021.^4,5^ From December 2020, England started to experience a rapid growth in covid-19 hospitalisations with 7-day rolling average increasing three-fold to middle January 2021. To control the spread and ‘Protect the NHS’ (The National Health Service), a localised three-tier system first introduced in October 2020 was re-introduced in December 2020, following a month-long second national lockdown in between.

During the pandemic, such non-pharmaceutical interventions (NPIs) made crucial contribution, particularly to alleviating hospital strains when effective treatments/vaccines were still widely inaccessible.^6,7^ Literature evaluating NPI effects focuses on viral transmission dynamics on national or global scales,^8–10^ whilst much of the available evidence on covid-19 hospitalisation derives from simulated predictions on observed socio-economic and demographic data with compartmental models, having made assumptions about clinical progression characteristics and transmission patterns and dynamics.^9,11,12^ Findings from these studies were pivotal in policymaking to combat the pandemic. However, neighbourhood-level interactions between hospitalisation, clinical progression, and transmission following localised NPIs could differ from national or global simulation assumptions.

We did an English literature search on quantifying localised NPI effects upon covid-19 health burdens and found seven relevant studies. One study predicted that physical distancing would have significantly reduced infections in Wuhan (China) by implementing return-to-work in early April 2020.^13^ Another study found transmission substantially reduced in Vo (Italy) after the community lockdown between 24^th^ February and 8^th^ March 2020.^14^ Two studies examined effects of localised restrictions in Chile during 2020: localised lockdowns between 21^st^ March and 4^th^ May reduced new cases in higher-but not lower-income areas;^15^ localised lockdowns between 3^rd^ March and 15^th^ June reduced transmission through strong modulation of the duration and indirect effects of neighbourhood interconnectedness.^16^ Two other studies found tiered restrictions effective in reducing transmissions, using reproduction number between 1^st^ July and 4^th^ November 2020 in the UK^17^ and case number between September 2020 and January 2021 in England^18^. Only one study examined hospitalisations under all tiers introduced in October 2020 in England and predicted a moderate reduction from then to 31^st^ March 2021.^19^

All seven studies have focused on either predicting future hospitalisation trends or effects on infections, which could be affected by prediction assumptions and levels of testing and case detection. Localised NPI effects on covid-19 hospitalisations in England, by comparison, have been insufficiently studied using observational methods. Evidence shows that covid-19 hospitalisation is less affected by case detection and a reliable indicator of the demand/burden of healthcare.^9,12,20^ Localised restrictions usually vary across neighbourhoods within countries that are at different phases of the pandemic with varied levels of resources/resilience.^14,16,18^ Gathering empirical evidence about local contexts is vital to deliver suitable health interventions to avoid overwhelming healthcare facilities and staff in places with high burdens.^9,12^ Such evidence is much needed to better gauge localised healthcare demand, particularly when observed hospitalisation data for neighbourhoods is available now in many countries including England.^21,22^

To fill our knowledge gap and lend support for the ongoing UK covid-19 inquiry, we therefore endeavour to analyse effects of Tier 3 restrictions introduced in December 2020 on covid-19 hospitalisations, compared to Tier 2 restrictions in this study. We further evaluate whether these intervention effects varied between small areas by level of deprivation and the prevalence of an Alpha variant (B.1.1.7).

## METHODS

### Patient and public involvement

Patients and public were not involved.

### Data and setting

We used Hospital Episode Statistics (HES) data provided by NHS Digital on weekly number of hospital admissions for covid-19 (International Classification of Diseases 10th edition: code U07.1 for confirmed infections, and U07.2 for suspected or probable infections by clinical or epidemiological diagnosis)^23^ in England between 4^th^ October 2020 and 21^st^ February 2021 as our outcome variable. They were aggregated to Middle Layer Super Output Areas (MSOA), a standard geographical unit in England covering around 20km^2^ and 8,000 residents on average.

We used measures of local area characteristics to adjust for potential confounders that could potentially affect vulnerability to hospitalisations and/or effectiveness of control measures. These included: overall score of the English Indices of Multiple Deprivation (IMD) 2019 - a composite indicator of socioeconomic disadvantage;^24,25^ total population and percentage of the over 70 using 2019’s mid-year estimates from the Office for National Statistics (ONS);^25^ proportion of Black, Asian, and Mixed ethnicity (BAME);^26^ population density from the ONS;^27^ and the proportion of people with previous admissions for chronic disease(s) (cardiovascular disease, diabetes, chronic kidney disease or chronic respiratory disease) during 2014-2018 to measure co-morbidities and clinical vulnerability.^25^ To account for pre-intervention LA-level accessibility differences to SARS-CoV-2 polymerase chain reaction (PCR) testing and the prevalence of B.1.1.7,^28^ we included the number of LA-level PCR tests per capita from the covid-19 dashboard and the proportion of LA-level positive tests with PCR S-gene test failure (SGTF) from Public Health England.

We then merged this time series of MSOA weekly data, area characteristics and linked LA data, with a dataset of LA-level tiered restrictions compiled by The Open Data Institute.^29^

### Intervention

Tier 3 intervention was implemented on 7^th^ December 2020 (Monday) by many areas, though officially announced on 2^nd^ December 2020. We based our analysis on this initial tier allocation, as tiers of most MSOAs remained unchanged except Kent’s MSOAs switched to a new ‘Tier 4’ on 20^th^ December 2020. Analysing the initial tier allocation could offer a more conservative estimate of effect size and avoid biases of selecting places based on subsequent tier transitions, resembling a trial where tier allocation itself is influenced by the effectiveness of restrictions.

We chose concurrent Tier 2 as the ‘control’ to our intervention. Although both tiers prohibited indoor mixing between households, Tier 3 was more restrictive than Tier 2. Meeting with people outside one’s own household in private gardens was not permitted in Tier 3 areas, whilst such meetings with up to six people were allowed in Tier 2 areas. Pubs and restaurants were closed in Tier 3 areas except for providing takeaway food, but remained open in Tier 2 areas if they served ‘substantial meals’ constituting main courses rather than just snacks.

### Analysis

We applied the synthetic control method for microdata developed by Robbins et al..^30,31^ The synthetic control group is constructed using a weighted combination of non-intervention areas. The intervention effect is estimated by comparing the post-intervention trend in outcome between the intervention and the synthetic control group.^32^

Accounting for covid-19’s incubation and clinical progression we assumed a two-week lag from the intervention onset (7^th^ December 2020) to observed impacts on hospitalisation (20^th^ December 2020).^33–35^ We measured covid-19 hospitalisation changes in the intervention and the synthetic control group, 12 weeks before and 9 weeks after 20^th^ December 2020 (between 4^th^ October 2020 and 21^st^ February 2021). We used this extended period to understand the extent to which Tier 3 effects (compared to Tier 2) were sustained.

To create the synthetic control group, we derived calibration weights to match Tier 2 and Tier 3 MSOAs over the pre-intervention period based on previously described eight local area characteristics and baseline hospitalisation levels. The calibrated weights met two conditions: the control group’s weighted average for each the local area characteristics equalled that of the intervention areas, and the total number of covid-19 hospitalisations in the control group matched that of the intervention areas for each pre-intervention week, minimising potential differences in unobserved characteristics associated with covid-19 hospitalisation rates or Tier 3 allocation.

We then estimated the Average Treatment Effect for the Treated (ATT) as the difference in cumulative number of covid-19 hospital admissions in the post-intervention period in the intervention areas, compared to the (weighted) cumulative number of admissions in the synthetic control group. To estimate the sampling distribution of the intervention effect and calculate permuted p-values and 95% confidence intervals, we generated 250 placebo iterations randomly allocating Tier 2 MSOAs to the intervention group, with sufficiently stabilised and converged distribution of outcomes.^31^

Tier 3 restrictions coincided with widespread community testing (piloted in Liverpool), which had a relatively large effect on hospitalisation.^32^ We thus excluded 342 MSOAs from our analysis (200 in Liverpool City Region, and 142 with higher than 1 per 100 population mean weekly lateral flow test (LFT) rate during community testing rollout between 6^th^ November 2020 and 2^nd^ January 2021: Supplement 1). This left 2809 Tier 3 MSOAs as the intervention group, whilst the synthetic control was constructed from 3481 Tier 2 MSOAs.

We further conducted interaction analyses to investigate whether the intervention effect varied between MSOAs by deprivation (IMD terciles) and the prevalence of B.1.1.7 (median). We re-ran the weighting algorithm, stratifying by terciles or median of the relevant variables. The ATT was estimated using stratified calibration weights in a weighted Poisson model with a log link function alongside an interaction term between the stratified variable and the intervention indicator.

### Sensitivity analysis

To test potential Tier 4 impacts, we experimented removing Kent’s MSOAs from our analysis (Supplement 2). To assess appropriateness of our research design in adjusting impacts of community testing, we further carried out a sensitivity test by including 342 MSOAs with higher LFT testing rates (Supplement 3). To check the robustness of our approach in potential spatial spill-over effects among neighbouring Tier 2 and 3 areas, we re-ran the main analysis by excluding the Tier 2 MSOA areas with population weighted centroids located within 20 km of Tier 3 areas (Supplement 4).

All analysis was performed using R version 4.3.2 and the Microsynth package.^30^

## RESULTS

### Descriptive analysis

We identified 2809 Tier 3 MSOAs as the intervention group and constructed the synthetic control group from the 3481 Tier 2 MSOAs: the brighter the colour, the lower the weights, and vice versa (Figure 1). Figure SF4 in the supplement shows location of synthetic control and intervention areas by Local Authorities.

**Figure 1.**
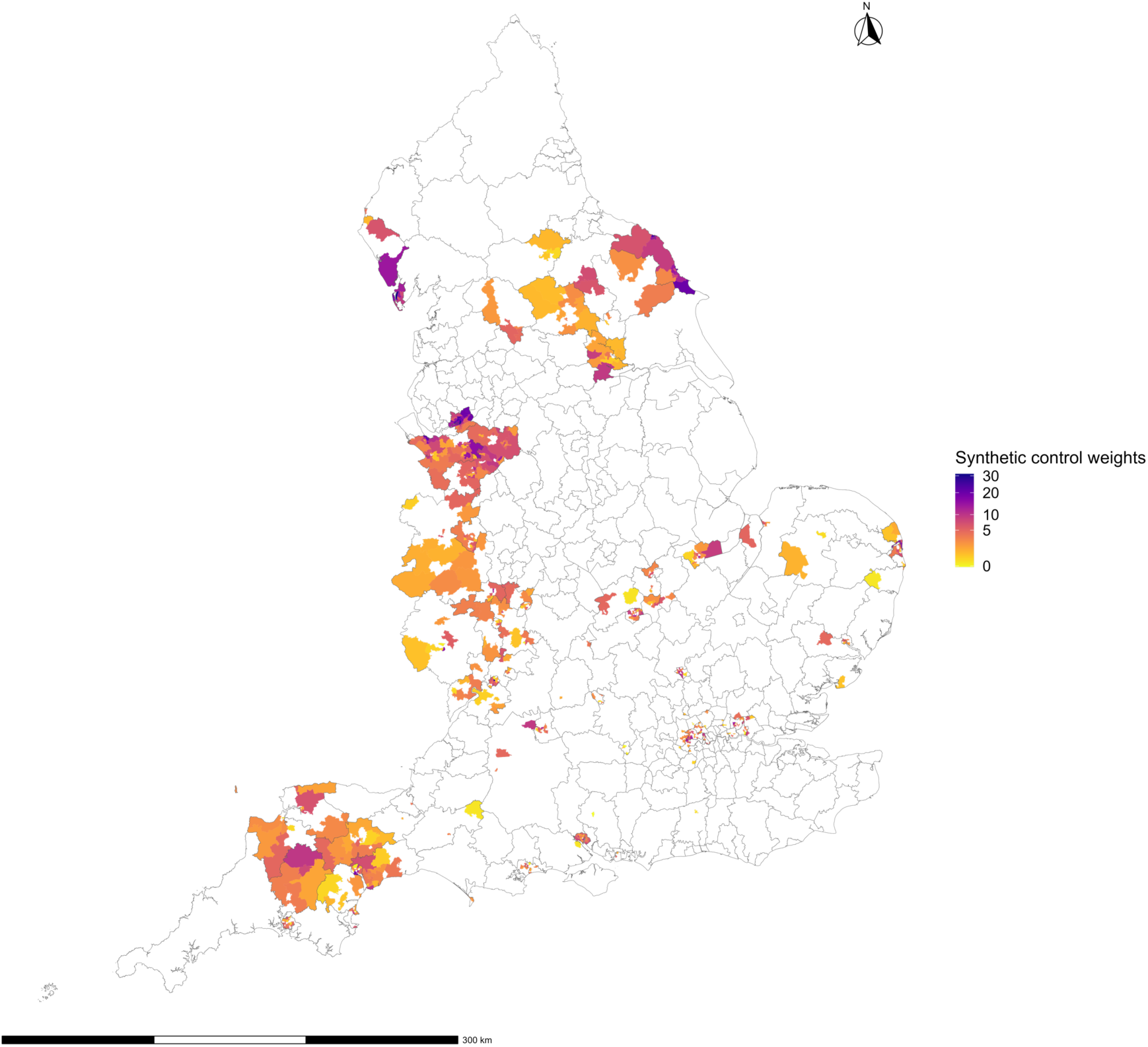
Weighting of Tier 2 Middle Layer Super Output Areas (MSOAs) that were used to construct synthetic control group at the intervention time point.

Table 1 presents summary statistics for the Tier 3 areas and the Tier 2 MSOAs from which the synthetic control group was constructed. Intervention (Tier 3) MSOAs have higher levels of deprivation, lower population density, higher proportion of the population with historical admissions for chronic disease(s), lower proportion of the BAME population, more PCR tests per capita, and lower proportion of SGTF. There was no difference in proportion of the population over 70. In the 12 weeks before the effects of Tier 3 upon hospitalisation kicking in on 20^th^ December 2020, Tier 3 areas had a higher covid-19 admission rate than Tier 2 MSOAs. As the matching algorithm achieved an exact match between the intervention and synthetic control areas, the weighted average of each variable in Table 1 would become identical for both groups after applying the calibrated weights.

**Table 1.** Comparison between Tier 3 areas and the MSOAs in the rest of England used to construct the synthetic control group (excluding those within Liverpool City Region or with a high LFT testing rate).

|  | **Tier 2 MSOAs used to construct the synthetic control** | **Tier 3 areas** |
| --- | --- | --- |
| **Index of Multiple Deprivation score** | 18 | 26 |
| **Population density – people per hectare** | 42 | 30 |
| **% of population aged 70+ years** | 14 | 14 |
| **% Black Asian and Minority Ethnic (BAME)** | 16 | 12 |
| **% S-gene target failure (SGTF) in routine PCR** | 43 | 23 |
| **Average PCR tests per 100,000 per week in 12 weeks before intervention introduced** | 2362 | 2786 |
| **% of population with at least 1 admission for chronic disease(s)** | 19 | 21 |
| **Average hospital admissions per 100,000 per week for covid-19 in 5 weeks before intervention introduced** | 7 | 14 |
| **Total population** | 29369663 | 22960484 |
| **Number of MSOAs** | 3481 | 2809 |

### Statistical analysis

We visualised the trend in the average weekly covid-19 hospital admission rates in the intervention and synthetic control areas 12 weeks before and 9 weeks after Tier 3 taking effect on 20^th^ December 2020 (Figure 2). These trends were identical in both groups prior to the intervention (4^th^ October to 19^th^ December 2020). In October and the first half of November the hospitalisation rates were increasing before effects of the second national lockdown upon hospitalisation were felt (assuming the two-week lag to observe). They then started to fall until mid-December. With the re-introduction of Tier 3 restrictions along with their effects upon hospitalisation starting two weeks later in December 2020, hospitalisation rates were increasing again until mid-January 2021. This increase however was slower in the Tier 3 areas compared to the synthetic control. From 18^th^ January until 21^st^ February 2021, hospitalisation rates were rapidly falling likely because of the third national lockdown on 6^th^ January 2021. The lower trend in Tier 3 areas continued from late December 2020 to early February 2021, after which no differences were observed between the two groups.

**Figure 2.**
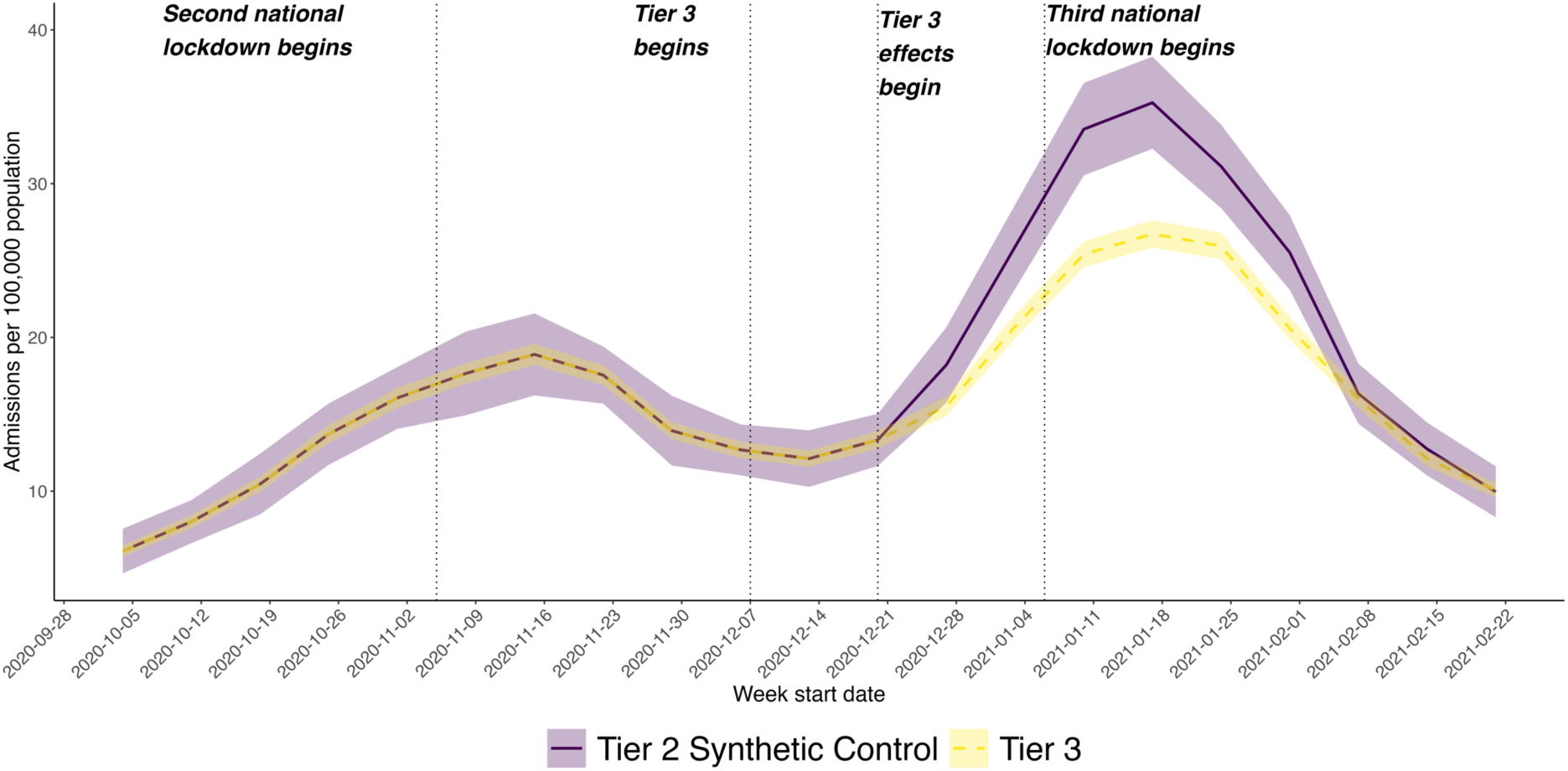
Trend in weekly covid-19 hospital admission rates in MSOAs in Tier 3 areas compared to a synthetic control group constructed from the weighted average of Tier 2 MSOAs after removing the effect of community testing. Dotted vertical lines represent start of the second national lockdown on 5^th^ November 2020, followed by the re-introduction of Tier 3 restrictions on 7^th^ December 2020 with a two-week’s lag on affecting hospital admissions from 20^th^ December 2020, before the third national lockdown on 6^th^ January 2021.

We then estimated the overall effect of Tier 3 restrictions to what would have been expected if Tier 2 restrictions had been applied on those areas (Table 2). Tier 3 restrictions implemented in December was associated with an average change of covid-19 hospital admissions of -17% (95% CI -21% to -13%; total change -8158, 95% CI -9981 to -6286), compared to the synthetic control.

**Table 2.** Results of the synthetic control analysis with a breakdown per level of deprivation (3 levels) and variant B.1.1.7 (binary following dichotomisation) – indicating the relative reduction in infections in Tier 3 areas compared to what would have been expected if Tier 2 restrictions had been applied. Although differences in effect are observed across levels of deprivation and prevalence of variant B.1.1.7 (indicated by S-gene target failure (SGTF) where quantitative reverse transcriptase PCR is used for covid-19 diagnosis), the interaction terms between level of deprivation and intervention, and between S-gene target failure and intervention were not statistically significant.

|  | Percentage change in admissions (95% CI) | Total change in admissions (95% CI) | P value | P value for interaction between Tier 3 intervention and levels of deprivation and prevalence of the variant respectively |
| --- | --- | --- | --- | --- |
| **All Tier 3** | -17% (-21% to -13%) | -8158 (-9981 to -6286) | <0.001 |  |
| Most affluent areas | -19% (-23% to -14%) | -1767 (-2261 to -1216) | <0.001 |  |
| Intermediate deprivation | -16% (-22% to -10%) | -1982 (-2742 to -1234) | <0.001 | 0.434 |
| Most deprived areas | -18% (-26% to -8%) | -4687 (-6756 to -2043) | <0.001 | 0.841 |
| Lower SGTF (2%-28%) | -17% (-27% to -7%) | -6326 (-10136 to 2553) | <0.001 |  |
| Higher SGTF (29%-85%) | -13% (-18% to -8%) | -1154 (-1599 to -670) | <0.001 | 0.379 |

In the subgroup analysis by deprivation, the effect size and direction were similar across all levels of deprivation suggesting that the benefits of regional tiers were observed across all levels of socioeconomic deprivation (Table 2). Although the effect was slightly greater in both the most deprived and affluent areas, given overlapping confidence intervals, there is no evidence to suggest that deprivation modifies the effects of regional tiers (and no evidence of widening inequalities).

We find evidence for differences in the estimated effect by the level of median proportion of B.1.1.7 cases. Tier 3 effects may have been greater in areas where the variant was less prevalent (-17%, 95% CIs -27% to -7%; total change -6326, 95% CI -10136 to 2553) than those of higher prevalence (-13%, 95% CIs -18% to -8%; total change -1154, 95% CI -1599 to -670). However, due to overlapping confident intervals and no statistically significant interaction effect, such observation cannot be robustly inferred.

### Sensitivity analysis

In sensitivity analysis we repeated the synthetic control models by removing the 182 MSOAs in Kent and found slightly smaller effects (Supplement 2). We also replicated the analysis without excluding the 342 MSOAs and found slightly larger effects (Supplement 3). We found almost identical effects in testing the potential spatial spill-over effects (Supplement 4).

## DISCUSSION

We found that Tier 3 restrictions implemented in December 2020 were associated with reductions in covid-19 hospitalisations in local communities (MSOAs) of England, compared to what would be the case if they had been put into Tier 2 restrictions instead. Tier 3 areas had 17% fewer covid-19 hospital admissions over the study period with the deviation in trends being observed two weeks post-implementation of the policy and quickly diverging from trends in Tier 2 areas. This effect was consistent across all levels of deprivation suggesting that that policy did not result in widening inequalities. The effect of regional tiers was potentially higher in areas where B.1.1.7 was less prevalent suggesting that their benefits may only be restricted to less infectious strains. Our study provides novel empirical evidence for the benefits of localised restrictions in managing covid-19 hospitalisations.^18^

### Strengths

We add to the evidence that localised restrictions on outdoor meeting and the hospitality sector in Tier 3 (compared to Tier 2) had an important role to play in managing covid-19 hospitalisations. Combined with our previous finding of regional tiers reducing SARS-Cov-2 transmission,^18^ our study here shows that these effects also extend to consequential covid-19 hospitalisation.^18^ As Tier 3 restrictions were more restrictive in outdoor meeting and the hospitality sector than Tier 2, we unfortunately cannot tell from our analysis whether either one, or a combination of both caused the observed effects. Previous evidence indicates that hospitality settings contribute to transmission^36,37^ but outdoor proximity has a low risk,^38^ which can lead potential reduction on consequential hospitalisations. No consistent evidence supports differentiated effects by levels of deprivation. Whilst national lockdowns required all people to work from home where possible, they may have had different effects across different socioeconomic groups as more deprived people were less likely to work from home.^39^ Additional restrictions on outdoor meeting and the hospitality sector in Tier 3 may have not had such differential effects by socioeconomic group, as transmission in outdoor settings and within the hospitality sector occurred at a similar level across different socioeconomic groups and thus the additional restrictions in Tier 3 reduced risks of consequential hospitalisations by similar amounts.^18^

### Limitations

Our study has limitations. Firstly, we assumed constant infection hospitalisation rate rover our relatively short study period. It could have, however, varied in England due to covid-19 variants that were more virulent but unaccounted for during our study. Current evidence does not associate the then variant B.1.1.7 with more severe disease suggesting our assumption was reasonable.^40^ Secondly, although we can match areas to ensure a good balance of potential confounding factors before Tier 3 causing effects, concurrent policy changes could bias the results. Community testing was one affecting transmission and the consequential hospitalisations then. Although we have sought to account for it, we made the adjustments assuming that the average effects of community testing on hospitalisation were proportional to the transmission levels in Tier 2 and Tier 3 areas in England, coupled with our earlier assumption of the constant infection hospitalisation rate. Kent went into Tier 4 in late December 2020, whilst results of the sensitivity test removing Kent from our analysis further confirmed the robustness of our main findings (Supplement 2). Thirdly, we were only able to use data on neighbourhoods, and thus unable to investigate how Tier 3 effects varied by individual or household characteristics.

## CONCLUSIONS

With increased access to vaccination and emergence of less deadly variants, many countries, including the UK, lifted covid-19 restrictions to help people get "back-to-normal". Globally, however, covid-19 has created large regional and national differences in related disease burden and healthcare needs. Localised restrictions suited to local contexts may remain needed to contain regional outbreaks and reduce pressures on local health systems. Concerns should be raised about the top-down and one-size-fits-all nationwide approaches, given that more deprived areas tend to be more affected economically, and have lower vaccination coverage, poorer healthcare facilities and higher disease burden. Our analysis indicates that tiered restrictions in outdoor gathering and in the hospitality sector are effective at moderately reducing hospitalisation and could be part of an effective strategy for reducing geographical differences in health burden among neighbourhoods as we exit from the pandemic. As the UK conducts a thorough inquiry into the handling of the pandemic, our research ensures a granular examination of tiers implemented at the community level, providing critical evidence not only for immediate response efforts but also for informing future public health policies and preparedness strategies.

### Contributors

BB and IB are the guarantors and accept full responsibility for the work and/or the conduct of the study, had access to the data, and controlled the decision to publish. BB and IB conceived the original idea, devised the project, the main conceptual ideas and proof outline. XZ conducted the literature review, collected the data, carried out the data analysis and drafted the manuscript. All authors contributed to the interpretation of data, provided critical feedback, and helped to shape the research manuscript and subsequent revision. The corresponding author attests that all listed authors meet authorship criteria and that no others meeting the criteria have been omitted.

## Data Availability

The small-area covid-19 hospital admissions data were made available by NHS Digital under data sharing agreement DARS-NIC-16656-D9B5T-v3.10 and are available through application to NHS Digital. All other data are publicly accessible, and code is available via the Liverpool City Region Civic Data Cooperative GitHub public repository https://github.com/civicdatacoop/tier3_covid_hospitalisation.

## Patient consent for publication

Not applicable.

## Ethics approval

Not applicable as this study only used anonymised and aggregated data.

## SUPPLEMENT

### Supplement 1: Community testing rates over time

**Figure SF1.**
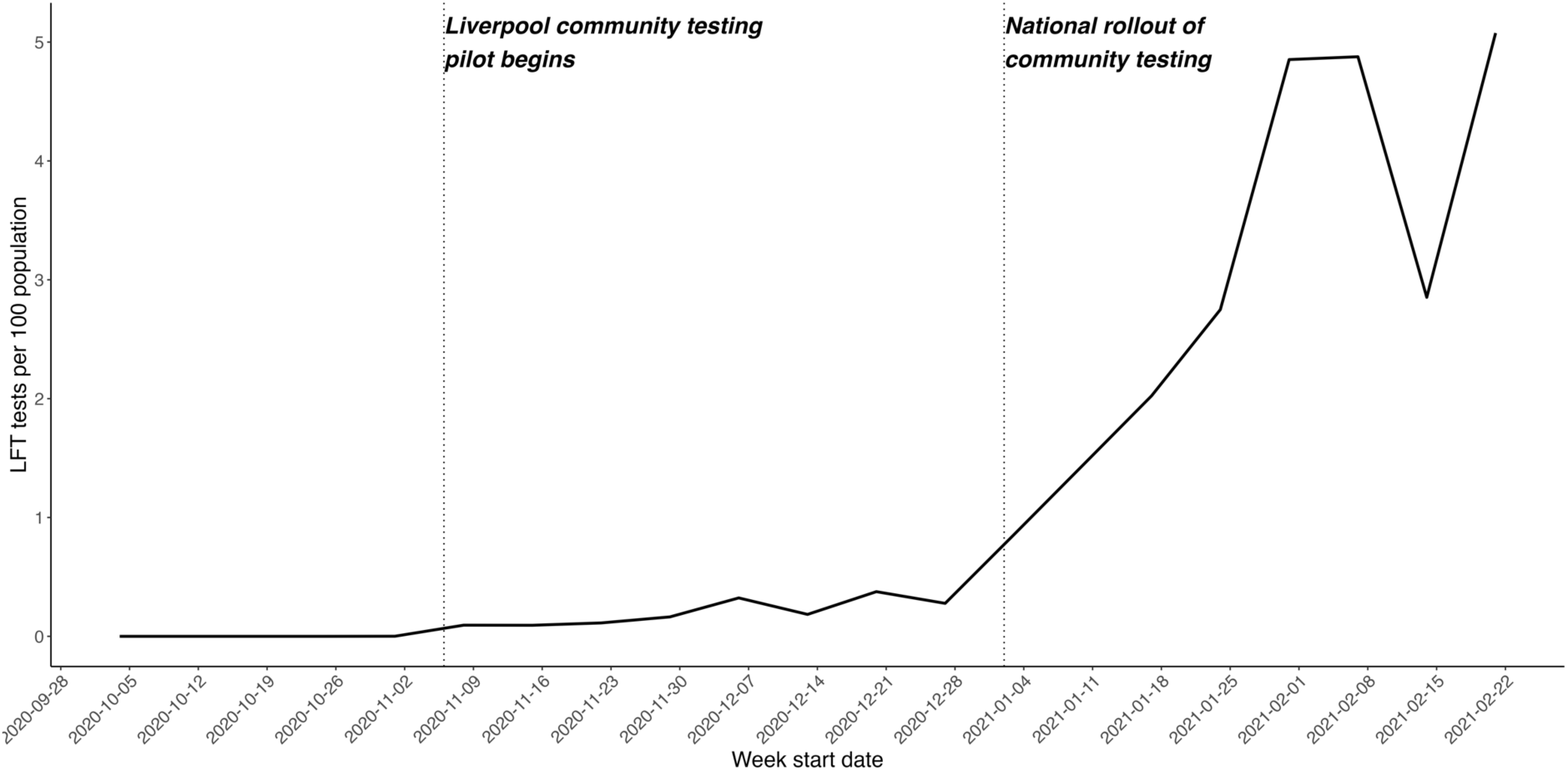
Trend of mean weekly SARS-CoV-2 antigen lateral flow tests (LFTs) per 100 population across Local Authorities in England between 4^th^ November 2020 and 21^st^ February 2022. Note: Dotted vertical line identifies onsets of community testing pilot in Liverpool and national rollout.

**Figure SF2.**
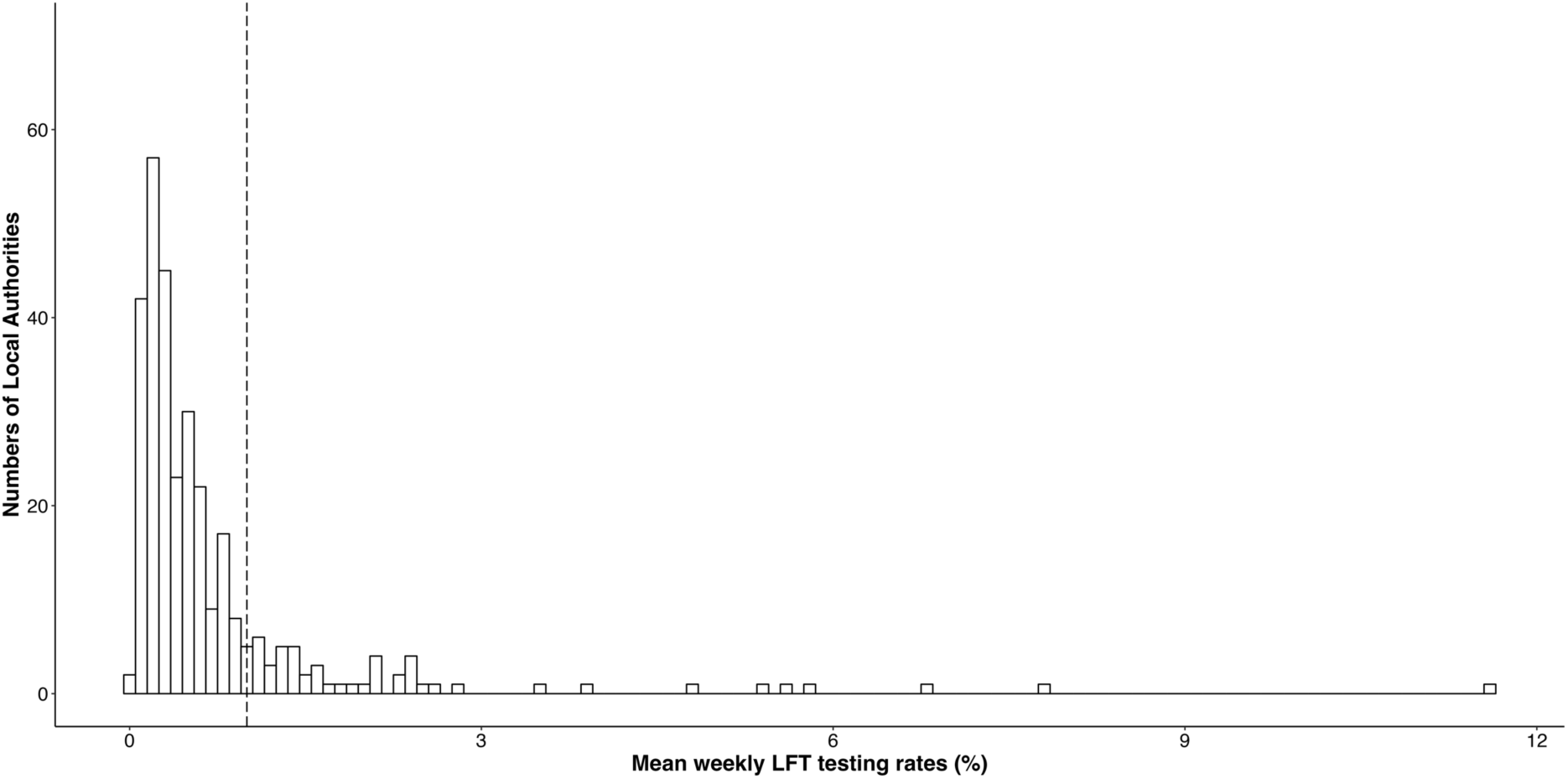
Distribution of mean weekly SARS-CoV-2 antigen lateral flow tests (LFTs) per 100 population across Local Authorities in England between 6^th^ November 2020 and 2^nd^ January 2021. Note: Dotted vertical line identifies the threshold of the mean LFT testing rate of 1 per 100 population per week that we used to exclude the 142 MSOAs with higher mean LFT rates from our analysis to minimise the potential impact of community testing on hospitalisation.

### Supplement 2: Sensitivity test of removing the MSOAs in Kent

**Table SF1.** Results of the synthetic control analysis after excluding the MSOAs in Kent – indicating the relative reduction in infections in Tier 3 areas compared to what would have been expected if Tier 2 restrictions had been applied.

|  | Percentage change in cases | 95% CI |  | p-value |
| --- | --- | --- | --- | --- |
|  |  | LCL | UCL |  |
| **All Tier 3** | -14% | -20% | -8% | <0.001 |

### Supplement 3: Sensitivity test of including the 200 MSOAs in Liverpool City Region and the 142 MSOAs with mean LFT rates above 1 per 100 population per week

**Table SF2.** Results of the synthetic control analysis by including the 200 MSOAs in Liverpool City Region and the 142 MSOAs with mean LFT rates above 1 per 100 population per week – indicating the relative reduction in infections in Tier 3 areas compared to what would have been expected if Tier 2 restrictions had been applied.

|  | Percentage change in cases | 95% CI |  | p-value |
| --- | --- | --- | --- | --- |
|  |  | LCL | UCL |  |
| **All Tier 3** | -19% | -23% | -16% | <0.001 |

### Supplement 4: Sensitivity tests of the spatial spill-over effect

The distance between MSOA areas is measured by the Euclidean distance of the population weighted centroids of MSOA areas (Data source is ONS Geography Open Data https://geoportal.statistics.gov.uk/datasets/b0a6d8a3dc5d4718b3fd62c548d60f81_0). When further excluding the 1,172 Tier 2 MSOA areas located within 20 km of Tier 3 areas (about 34% of the 3,481 eligible MSOAs), we found almost identical result (see Table SF3). This suggests that our results are robust and there probably have been very little spill-over effects. In other words, there is no evidence from our data that traveling from Tier 3 areas to neighbouring Tier 2 areas to take advantage of the less restrictive measures had made any contribution to hospitalisation for COVID-19 in neighbouring Tier 2 areas.

**Table SF3.** Results of the synthetic control analysis by excluding the 1,172 Tier 2 MSOA areas located within 20 km of Tier 3 areas – indicating the relative reduction in infections in Tier 3 areas compared to what would have been expected if Tier 2 restrictions had been applied.

|  | Percentage change in cases | 95% CI |  | p-value |
| --- | --- | --- | --- | --- |
|  |  | LCL | UCL |  |
| All Tier 3 | -16% | -21% | -10% | <0.001 |

**Figure SF3.**
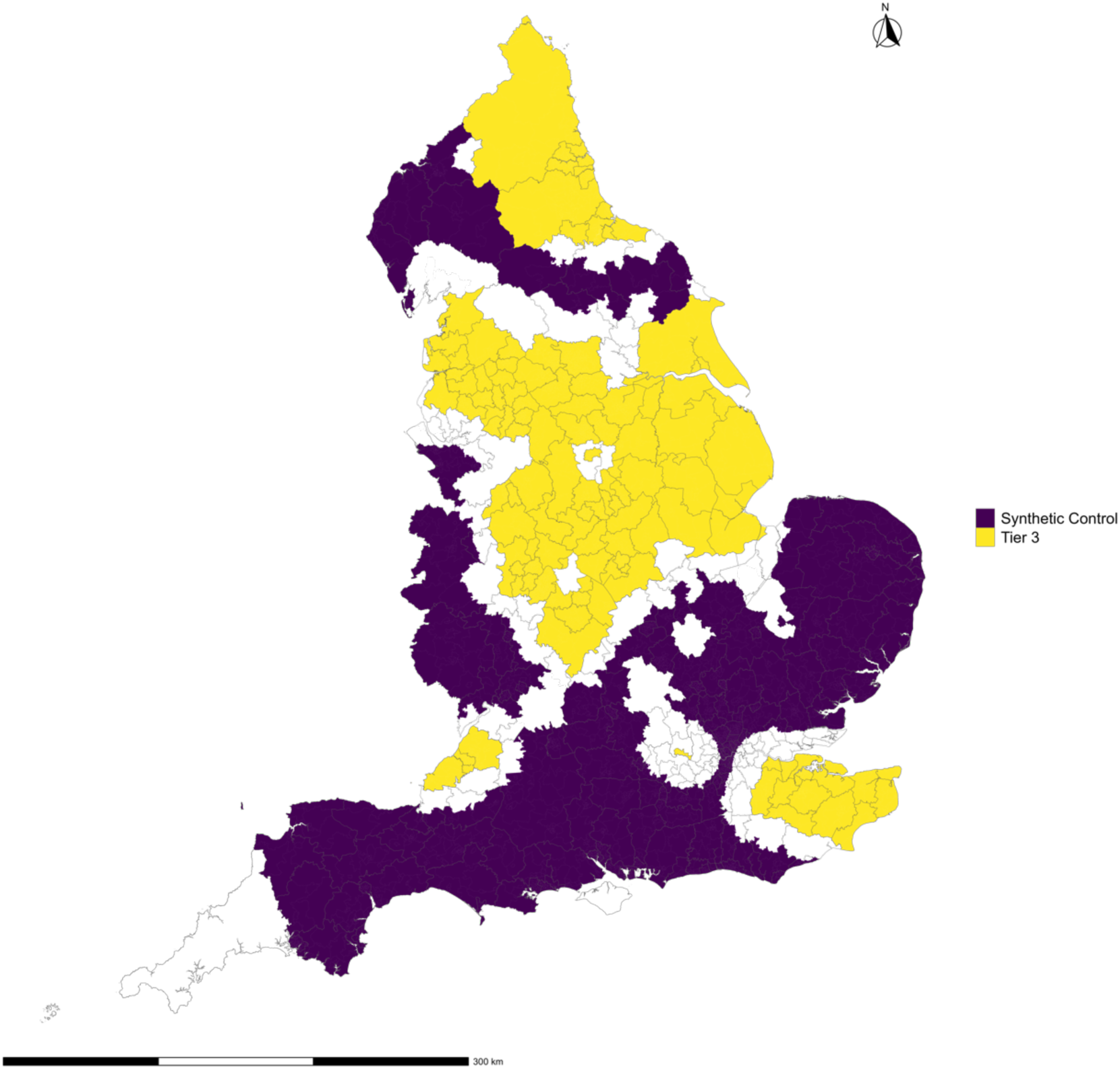
Location of synthetic control (yellow) and intervention (Tier 3) areas (purple) after excluding the 1172 Tier 2 MSOA areas located within 20 km of Tier 3 areas.

### Supplement 5: Location of Tier 2 areas used to construct synthetic control group

**Figure SF4.**
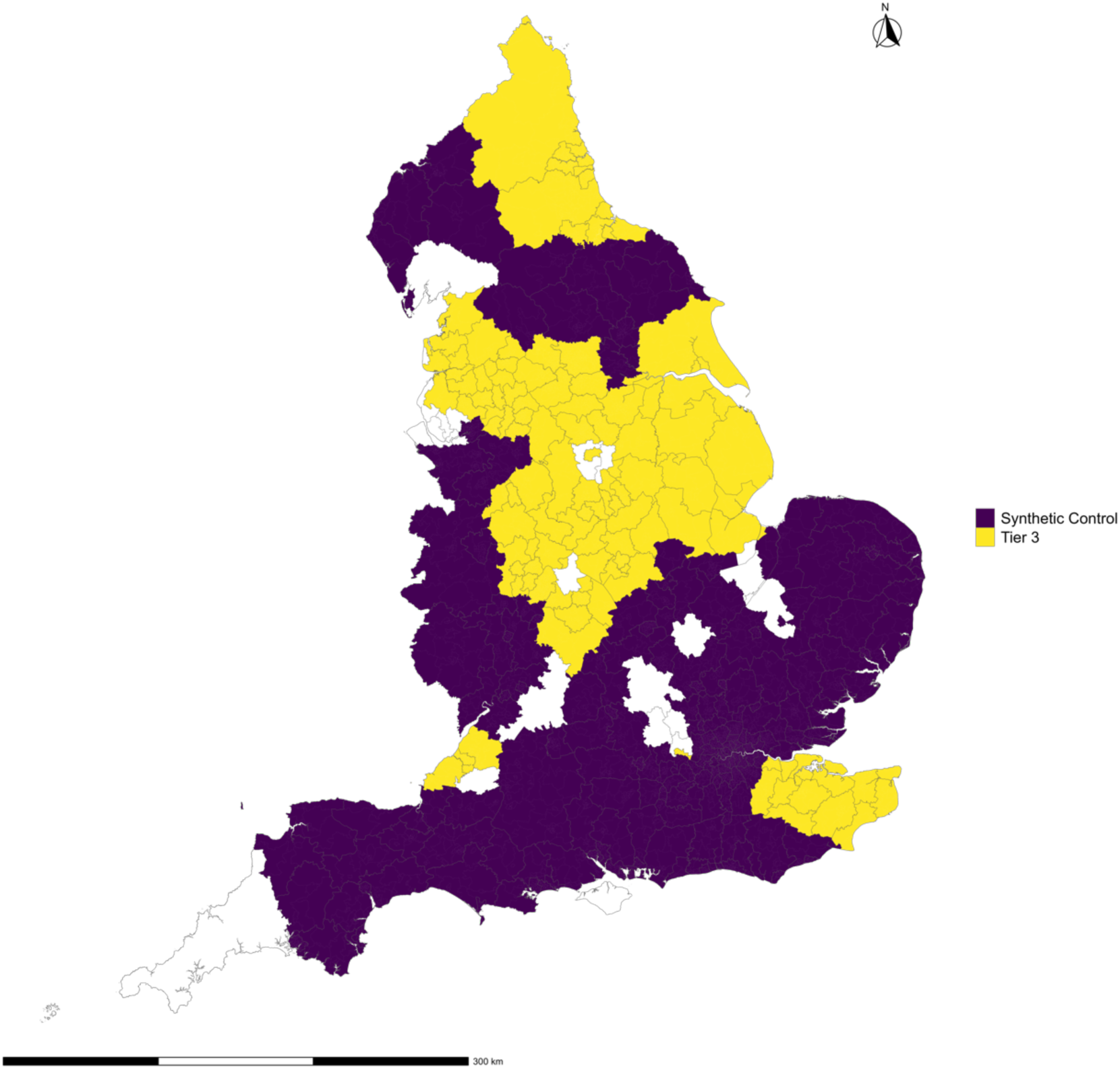
Location of synthetic control (purple; Tier 2) and intervention (Tier 3) areas (yellow) by Local Authorities.

